# Closing the gap: Solving complex medically relevant genes at scale

**DOI:** 10.1101/2024.03.14.24304179

**Authors:** Medhat Mahmoud, John Harting, Holly Corbitt, Xiao Chen, Shalini N. Jhangiani, Harsha Doddapaneni, Qingchang Meng, Tina Han, Christine Lambert, Siyuan Zhang, Primo Baybayan, Geoff Henno, Hua Shen, Jianhong Hu, Yi Han, Casey Riegler, Ginger Metcalf, Geoff Henno, Ivan K. Chinn, Michael A. Eberle, Sarah Kingan, Tim Farinholt, Claudia M.B. Carvalho, Richard A. Gibbs, Zev Kronenberg, Donna Muzny, Fritz J. Sedlazeck

## Abstract

Comprehending the mechanism behind human diseases with an established heritable component represents the forefront of personalized medicine. Nevertheless, numerous medically important genes are inaccurately represented in short-read sequencing data analysis due to their complexity and repetitiveness or the so-called ‘dark regions’ of the human genome. The advent of PacBio as a long-read platform has provided new insights, yet HiFi whole-genome sequencing (WGS) cost remains frequently prohibitive. We introduce a targeted sequencing and analysis framework, Twist Alliance Dark Genes Panel (TADGP), designed to offer phased variants across 389 medically important yet complex autosomal genes. We highlight TADGP accuracy across eleven control samples and compare it to WGS. This demonstrates that TADGP achieves variant calling accuracy comparable to HiFi-WGS data, but at a fraction of the cost. Thus, enabling scalability and broad applicability for studying rare diseases or complementing previously sequenced samples to gain insights into these complex genes.

TADGP revealed several candidate variants across all cases and provided insight into *LPA* diversity when tested on samples from rare disease and cardiovascular disease cohorts. In both cohorts, we identified novel variants affecting individual disease-associated genes (e.g., *IKZF1, KCNE1*). Nevertheless, the annotation of the variants across these 389 medically important genes remains challenging due to their underrepresentation in ClinVar and gnomAD. Consequently, we also offer an annotation resource to enhance the evaluation and prioritization of these variants.

Overall, we can demonstrate that TADGP offers a cost-efficient and scalable approach to routinely assess the dark regions of the human genome with clinical relevance.

## Introduction

Hybrid capture based DNA sequencing is a popular and cost-effective method to target genomic regions of interest. Exome sequencing (ES), for example, was heavily used to characterize variations in coding regions of the genome^1^. ES is a widely used tool in clinical genomics that has facilitated the identification of numerous deleterious mutations associated with diseases, including Robinow syndrome, Miller syndrome, and Fowler syndrome^2–4^. Additionally, ES has enabled the identification of causal mutations for conditions of previously unknown etiology^5,6^.

Despite its significant contributions to genetic research, ES has limitations that preclude it from definitively resolving all genetic diagnoses^7^. Over 50% of individuals suspected of having a genetic disorder remain undiagnosed even after thorough ES clinical assessment^8^. Moreover, approximately 98-99% of non-coding genomic regions are not covered by ES^9–11^. Additionally, ES prioritizes the detection of small variants (typically < 50bp) and copy number variants^12^, but frequently misses tandem repeats^13^, structural rearrangements^14^ and epigenetic modifications^7^. Most limitations stem from the use of short-read (SR) sequencing technologies, which can encounter mapping difficulties in regions with high sequence homology, such as *SMN1*, *CBS*, and *CYP21A2*^15^. Additionally, genes in GC/AG-rich regions^16^, those with extensive repetitive elements, or containing structural variants^14,17^ pose significant molecular and analysis challenges for ES^18,19^. Limitations of ES hinder our understanding of disease mechanisms and potentially lack of diagnosis.

Long read (LRs) sequencing methods can effectively capture structural rearrangements within single reads, simplifying analysis^14,20^ overcoming a key limitation of SR sequencing. The accuracy and length of long reads enable the more comprehensive characterization of ‘dark regions’ that are inaccessible to SR methodologies^21–23^. For instance, the *LPA* encompasses a large Variable Number Tandem Repeat (VNTR) known as KIV-2, which is a two-exon tandem duplication ranging from 1 to 40 copies^24^. The number of KIV-2 repeats correlates inversely with its lipoprotein protein A concentration as Lp(a), a predictor of cardiovascular diseases^25,26^. Resolving variants in KIV-2 is important but often hindered due to its complexity^27^. Additionally, genes with high polymorphism and repetitive elements, such as the Human Leukocyte Antigen (*HLA*) genes^28,29^ benefit from long read clinical *HLA* typing^30,31^. Similarly, *CYP2D6*, which is involved in the metabolism of >20% of common drugs, contains structural variations (SVs) that impact an individual’s response to certain medications^32,33^.

Using long reads, it is possible to phase variants in trans^34^, which is often required to accurately interpret functional consequences^35^ and establish genetic inheritance. For example, in *TPMT* gene, there are two variants impacting the gene’s function that are approximately 8 kb apart. If physically linked, in trans, these two variants can lead to Loss-of-function (LoF) of *TPMT*^36^. The consequences could lead to a complete knockout of *TPMT*, an important drug metabolizer, with potentially severe implications for patients^37^. The benefits of long reads make them attractive to large projects aimed at rare diseases and population diversity, such as the *All of Us* Research Program^20^, Genomics Research to Elucidate the Genetics of Rare diseases (GREGoR)^7^, and the Human Genome Structural Variation Consortium (HGSVC)^38^. Similarly, initiatives like Genome in a Bottle (GIAB) have leveraged long reads to improve benchmark datasets^39^. In the GIAB study, researchers investigated nearly 400 medically relevant genes that were previously inaccessible due to their highly repetitive and polymorphic nature, as well as the presence of segmental duplications and complex variants affecting these genes^39^.

In the GIAB benchmark study, they characterized 273 challenging medically relevant autosomal genes using a haplotype resolved whole-genome assembly. However, a limitation of LR technology is its higher cost compared to both SR whole-genome sequencing (SR-WGS) and ES^8^. This cost factor of long reads poses challenges for small labs, projects aiming for larger cohort sizes, or those operating in clinical settings, making it difficult to adopt long-read whole-genome sequencing (LR-WGS) as an initial screening approach.

We address the limitations of long read costs by developing a capture panel that targets ‘dark’ genomic regions, which are medically relevant, but difficult to resolve using ES or SR-WGS alone. We present the Twist Alliance Dark Genes Panel (TADGP) together with an analytical workflow. The panel design captures 389 complete autosomal genes, including both intronic and exonic regions, spanning a total of 22.20 megabase pairs (Mbp). The selection of the 389 genes was based on our prior efforts aimed at resolving challenging, medically relevant genes (CMRG)^39^. We have added three additional genes given their clinical importance, namely, *GBA* (associated with Gaucher disease), *CYP2D6* (linked to variation in drug metabolic rates), and *SBDS* (implicated in Shwachman-Diamond syndrome)^40–43^. The TADGP leverages the capabilities of PacBio HiFi reads, resolving repetitive and complex genes. TAGDP identifies and phases SNVs, Indels (<50bp) and SVs (≥50pb) with high accuracy. The analysis is self-contained within a workflow which combines read mapping and assembly techniques to generate the best possible variant calls. The workflow reports a comprehensive list of variant classes, including pharmacological impact where appropriate. We accurately identified variants within these 389 complex genes, achieving a high F-score of 97.00% for SNVs and 93.50% for SVs. The TADGP provides equivalent variant calling accuracy when compared to LR-WGS at a fraction of the cost. We illustrated the utility of the panel across cell lines and patient samples, encompassing cardiovascular and rare disease cases. This panel is a cost effective solution for routine examination of these complex genes with high accuracy.

## Results

### Capture design and variant characterization approach

We developed a capture panel and analysis workflow for HiFi long read data called the TADGP, targeting 389 complex clinically relevant genes (listed in **Table S1**). These genes are only fully characterized using long reads, which are required for accurate variant calling^20,39^. We extended the 386 CMRG gene for the GRCh38 list to include *GBA, CYP2D6*, and *SBDS*. Furthermore, our gene panel encompasses five clinically actionable genes (i.e., variants that need to be reported if they impact these genes based on ACMG guidelines) associated with a diverse range of medical conditions^44^, as detailed in **Supplementary Table S2**. TAGDP genes are linked to neurological disorders (e.g., *GBA*-associated Gaucher disease), cardiovascular diseases (e.g., *LPA*-related lipoprotein(a) dyslipidemia), and cancer (e.g., *SIGLEC16*-positive myeloid neoplasms, *ESRRA*-driven thyroid and endometrial cancers), as shown in **Figure 1a**. As an example, 21.6% of the genes are associated with neurological diseases. TADGP encompasses a total of 22.20 Mbp (introns and exons) with 3 kb extensions up and downstream of each gene. Out of the 389 genes, four genes—*LCE3B*, *SNORD64*, *DUX4L1*, and *TRBV9*— lacked coverage due to the absence of probes. Overall 25% of the 389 genes contain 1,879 pathogenic ClinVar variants (see methods). We posit that the remaining 75% of the genes in the panel lack ClinVar variants due to difficulty of assessment stemming from their inherent complexity^39^. We anticipate that TADGP will facilitate additional discoveries linking genes to phenotypes. The number of reported ClinVar variants significantly varies across the genes, with *GBA* exhibiting the highest number of variants (**Supplementary Figure 1**). We compared the percentage of GC/AG content per gene with the average gene coverage obtained from using TADGP **Figure 1b and Supplementary Figure 2**. The TADGP panel has the highest normalized coverage among all methods (**Figure 1c**), indicating that the GC% has a minimal, but significant impact on the panel’s coverage performance (p-value 9.16e-6, correlation: −0.22). Additionally, there is a negligible correlation between AG% and average coverage (p-value 0.37, correlation −0.04).

**Figure 1.**
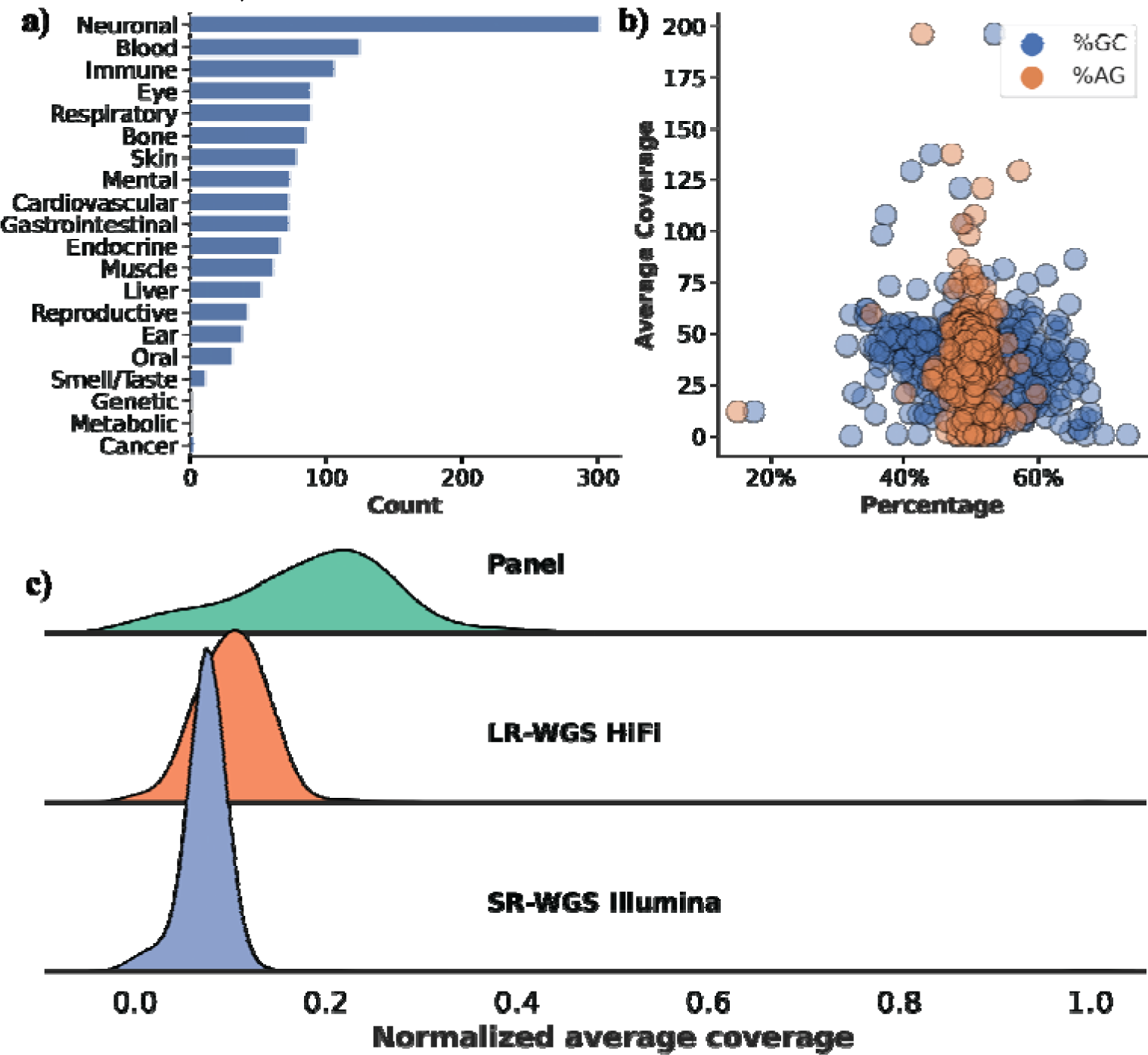
Characteristics of the 389 TADGP genes and their linked diseases. **a)** Gene count per disease, the X-axis is the count of genes associated with each disease, and the Y-axis is the disease category. **b)** GC% (in blue) and AG% (in orange) per gene vs. average gene coverage for sample HG002. The GC% affecting HiFi coverage is minimal, while AG% has no impact. **c)** Normalized average gene coverage between the panel (green), LR-WGS HiFi (orange), and SR-WGS Illumina (blue). TADGP has the highest normalized average coverage.

The computational workflow we developed is shown in **Figure 2**, which enables the characterization of variants across these 389 challenging genes. The output of the TADGP workflow includes phased SNVs, indels, SVs, pharmacogenomics clinical annotation, and *CYP2D6* allele diplotypes accurately assigning. The workflow starts with read alignment to a modified version of the GRCh38 reference genome as proposed by Behera *et al*^45^. The adapted reference mitigates issues in collapsed and duplicated regions. Next, small variants (SNVs and indels) are called with DeepVariant^46^ and structural variants are called with Sniffles2^47^. We included an assembly approach for 79 genes, due to their complexity. The supplementary section “Impact of TADGP Assembly over Mapping” summarizes the selection of these genes for the *de novo* assembly approach based on the lacking variant calling performance (**Supplementary Table S3)**.

**Figure 2.**
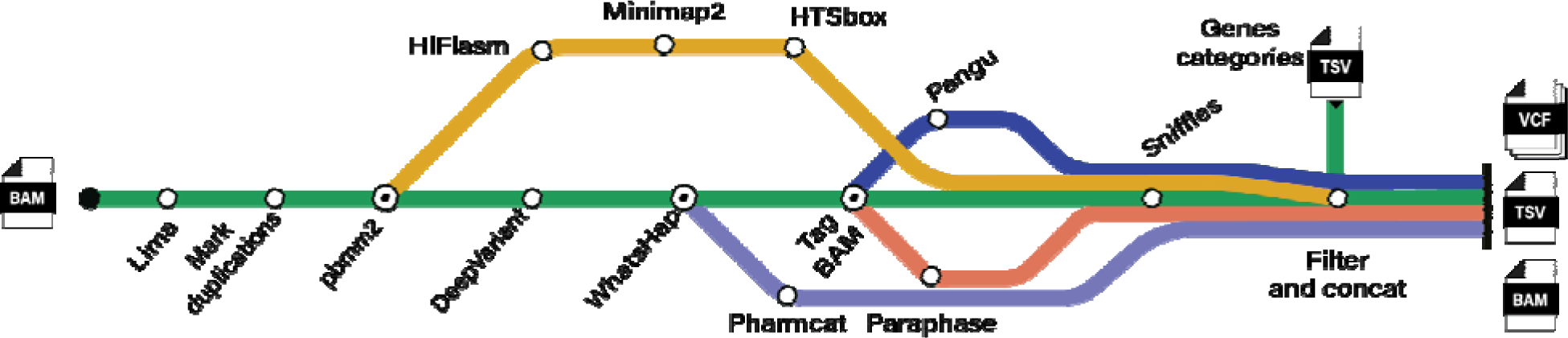
A schematic representation of the bioinformatic workflow implemented for characterizing complex medical-relevant genes. The mapping approach is shown in a green line, while the assembly process is shown with a yellow line. The tools used in the workflow are shown with different color bands, with Pharmcat in purple, Pangu in blue, and Paraphase in orange. Intermediate steps are marked with open circles, whereas endpoints are annotated with a dot. The workflow is contained within a snakemake workflow.

Our workflow which enables parallel analysis of multiple samples, implements several new bioinformatic tools specifically designed to genotype paralogous alleles including, the PacBio star-typer (Pangu)^48^ for *CYP2D6* and Paraphase^49^ for other paralogous genes such as *SMN1/SMN2, NCF1/NCF1B/NCF1C, PMS2* and *CFC1/CFC1B*. Lastly, the workflow consolidates all variant calls into standardized VCF files per sample (see methods). The workflow is publicly available on GitHub https://github.com/PacificBiosciences/HiFiTargetEnrichment under a BSD 3-Clause Clear license.

### Benchmarking TADGP on GIAB HG002

To validate our capture panel and bioinformatic workflow, we assessed the variant calling performance using the NIST Genome in a Bottle standard, HG002^39^. We sequenced three replicates of HG002 across three different sequencing runs (using Revio SMRT cells) to assess reproducibility. We obtained between 402,663 to 967,702 on-target reads for HG002 across three replicates, with a median read length of 5,244-5,647 bp and N50 size of 5,444-6,404 bp. The alignment rate ranged from 89.60% to 98.89%, and the median read quality score was Q35-36 (**Supplementary Table S4**). We measured the on-target gene coverage and the adequacy of coverage at individual base pair positions. The three technical replicates had an on-target rate between 49.69-54.90% (**Supplementary Table S5)**. We achieved a high average gene coverage from 38.34x to 99.75x. We measured the number of bases that exceeded a coverage of 8x, which we consider the minimum threshold for confidently identifying SNVs and indels. Based on our criterion, we found between 93.58% and 96.46% on-target gene bases that surpassed the 8x coverage threshold at MAPQ>=20. Even when increasing the threshold to 20x, we still observed between 86.55% and 93.63% of bases having >20x coverage across the 22.20 Mbp panel. We found that *OR4F5* is not covered in HG002 across all three replicates. For the first two HG002 replicates *DRD4*, *KISS1R*, and *TAS2R45* genes were not covered and *IGKV1-5* also was not covered in the second sample (**Supplementary Table S5).** The TADGP further captures the pseudogenes if present. To quantify this, we measured the coverage across pseudogenes for HG002. We identified 28 pseudogene copies across the panel design, showing an average coverage of 56.73x (**Supplementary Table S5)** (see methods). This is illustrated by *GBA* (49.77x coverage) in contrast to the *GBAP1* pseudogene copy with 16.90x coverage.

We next compared the TADGP to LR-WGS (30x PacBio Revio HiFi) and Illumina SR-WGS (32.44x). Across the 389 target genes, the normalized average coverage of the panel was 2.5-fold higher when compared to LR-WGS (**Figure 1c**). We used the CMRG benchmark dataset that consists of 21,232 SNV and indel calls (i.e. small variants <50bp), along with 217 SVs (>50bp) to benchmark the TADGP performance. When considering substitution and indel F-scores for the TADGP, LR-WGS, and SR-WGS were 94.6% (1,071 missed variants), 97.16% (390 missed variants), and 94.2% (1,264 missed variants), respectively (**Supplementary Figure 3**). The concordance of variants among the three technologies, including three panel replicates, was high (77.80%) (**Figure 3a**). The TADGP identified 963 small variants that were not captured by SR-WGS. Out of these, the panel uniquely identified 182 small variants (0.70%) not detected by either LR-WGS and SR-WGS. On the other hand, LR-WGS has 723 and SR-WGS has 717 falsely identified small variants. TADGP on the other side reduced this number to 83 falsely identified small variants. This suggests that the TADGP method offers advantages in specific genomic regions, enabling it to effectively address challenges that may be encountered with other approaches.

**Figure 3.**
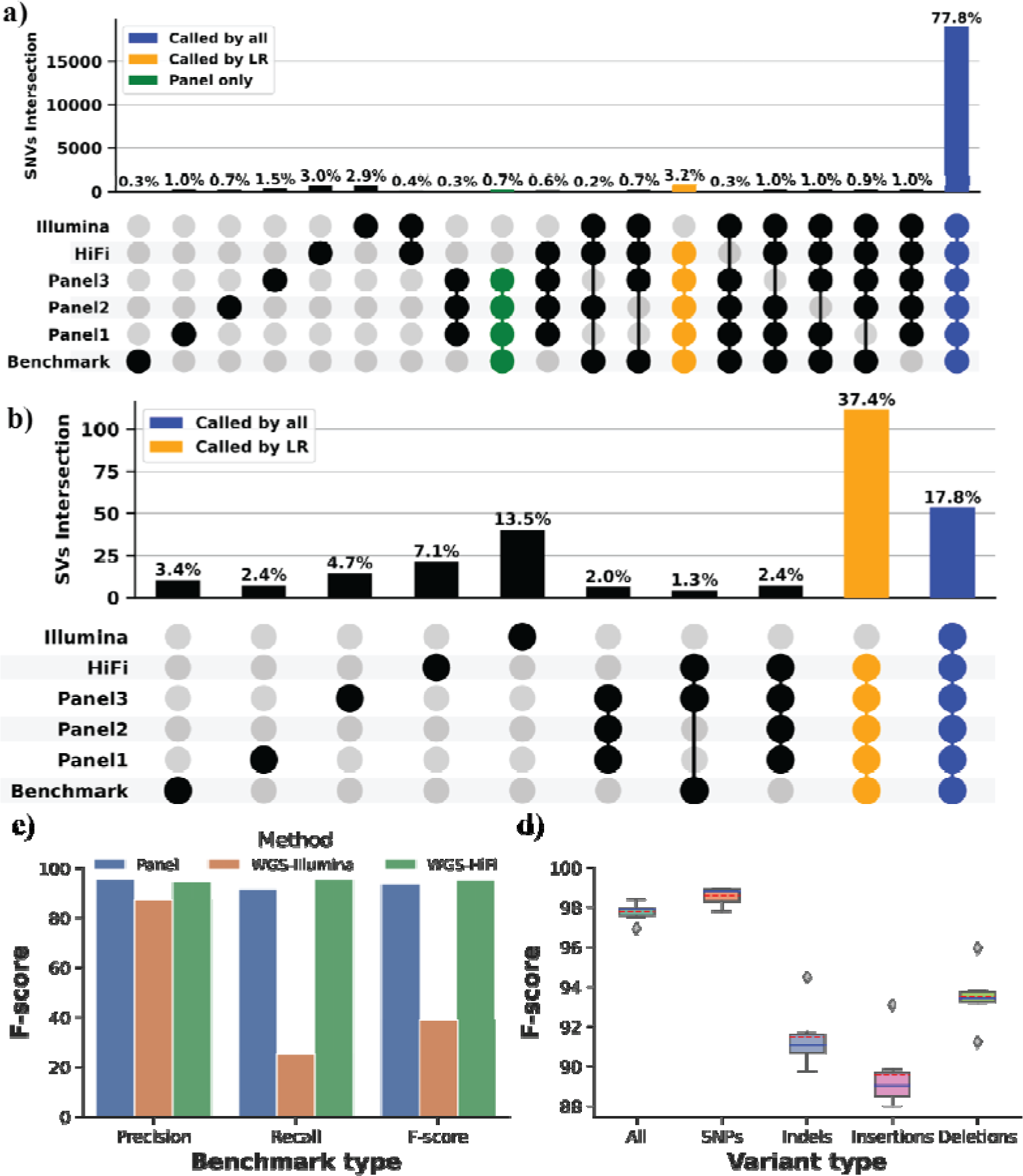
Performance assessment across samples HG001 to HG007. **a)** Comparison of unique and shared variants (SNVs/Indels) represented by Upset: TADGP (three replicate), HiFi LR-WGS, and Illumina SR-WGS for sample HG002. The blue column signifies the concordance observed among all datasets, the yellow column represents variants that were not called using Illumina dataset, and the green column highlights variants exclusive to the panel that are not present in either HiFi LR-WGS and Illumina SR-WGS. **b)** Comparison of unique and shared SVs represented by Upset. In the plot, the blue color represents SVs that are shared across all datasets, while the yellow color represents SVs that are absent when using the SR-WGS Illumina dataset. **c)** Comparison of SVs across the panel (Blue), WGS-illumina (Orange), and WGS-HiFi (Green), on the X-axis is the Precision, Recall, and F-score and the percentage on the Y-axis. **d)** A boxplot displays the F-score for SNVs and indels, for samples HG001, 3, 4, 5, 6 and 7 with the data split into five categories: All (SNPs and Indels), SNPs (substitutions only), Indels (insertions and deletions), insertions only, and deletions only. The Y-axis represents the F-score percentage, while the boxplot depicts the median with a solid blue line and the mean with a dashed red line.

We compared SVs performance amongst the TADGP, SR-WGS, and LR-WGS. The panel exhibited an overall F-score of 93.5%, similar to what we achieved using LR-WGS (94.99%), with a recall of 91.67% and a precision of 95.41%. For SR-WGS, the F-score was 38.85%, due to the missing SV calls (**Supplementary Figure 4**). Overall 18/217 SVs were missed by TADGP, out of these 11 were missed due to insufficient depth of coverage (**Supplementary Table S6 shows the coverage per SV and the SV type)** and TADGP only missed 3/14 SVs that intersect exons. Our concordance measures reveal that 127/217 of the SVs can only be detected using long reads, either from TADGP or LR-WGS, and remain undetected by SR-WGS (**Figure 2b**), from those 6/217 detected only by the TADGP. Furthermore, TADGP had six false positive SV versus 40 from Illumina and 21 from PacBio HiFi-WGS (**Figure 3b**).

Among the 389 genes covered by our panel, 116 genes were excluded by GIAB for their curated benchmark due to their complexity^39^. To benchmark the performance of TADGP, we utilized an existing assembly from the HPRC project to infer variants (**see Supplementary Table S7**) see methods. Over these 116 genes the panel achieved an F-score of 68.00% for SNVs and indels, and 68.40% for SVs. LR-WGS demonstrated a 70.89% F-score for SNVs and indels, while exhibiting a reduced performance of 56.92% for SVs. SR-WGS resulted in an even lower F1 score, achieving 64.86% F-score for SNVs and indels, and 13.18% for SVs (**Supplementary Figures 5-6**). We suspect that this subset of genes results in lower F-scores across technologies due to their inherent complexities in these regions and the lack of vetted benchmark data; they highlight that the panel performs well compared to WGS approaches from PacBio and Illumina. We also assessed the accuracy of Paraphase calls for *SMN1*/*SMN2*, *NCF1/NCF1B/NCF1C*, *PMS2/PMS2CL,* and *CFC1*/*CFC1B* against the T2T HG002 assembly. The F-score for SNVs and indels is 96.3% excluding the homopolymer regions.

Next, we compared the phasing performance between the TADGP and LR-WGS. The TADGP completely phased 63 genes into one phase block, while LR-WGS phased 111 genes. Furthermore, TADGP was able to phase 85.21% of the heterozygous variants per gene compared to 98.57% for LR-WGS (**Supplementary Figures 6 and 7**). The difference in performance between the TADGP and LR-WGS can be explained by the longer insert size of LR-WGS.

To increase the throughput of our assay, we have relied on multiplexing samples on a single Revio SMRT Cell. Our current run has been set up for 12 samples per Revio SMRT cell, which was conservative and yielded high coverage (38.34x to 99.75x) per sample. We subsampled (down to 10%) the coverage and assessed variant calling performance on HG002 data. As expected, the average coverage across the genes declined, but did not dramatically impact the variant calling performance (**Supplementary Figure 8**). Only at 10% of the original coverage, we have identified 15 uncovered genes. We further observed similar performance (F-score) across SNVs, Indels, and SVs (**Figure 4a**). Even on a per gene measurement (**Figure 4b**) we do not observe a large impact on variant calling. A reduction in accuracy is only observed at 10% coverage (10.92x average gene coverage). This goes in hand with a reduction of the F-score from 94.81% at the original coverage to 82.71% at the reduced 10% coverage level. Similarly, for SVs, only in the 10% titration we observed a reduced F-score from 87.57% to 83.99% (**Figure 4a**). Thus, TADGP could run up to 48 samples per PacBio Revio SMART cell which would further reduce the costs per sample and improve the throughput, without sacrificing accuracy.

**Figure 4.**
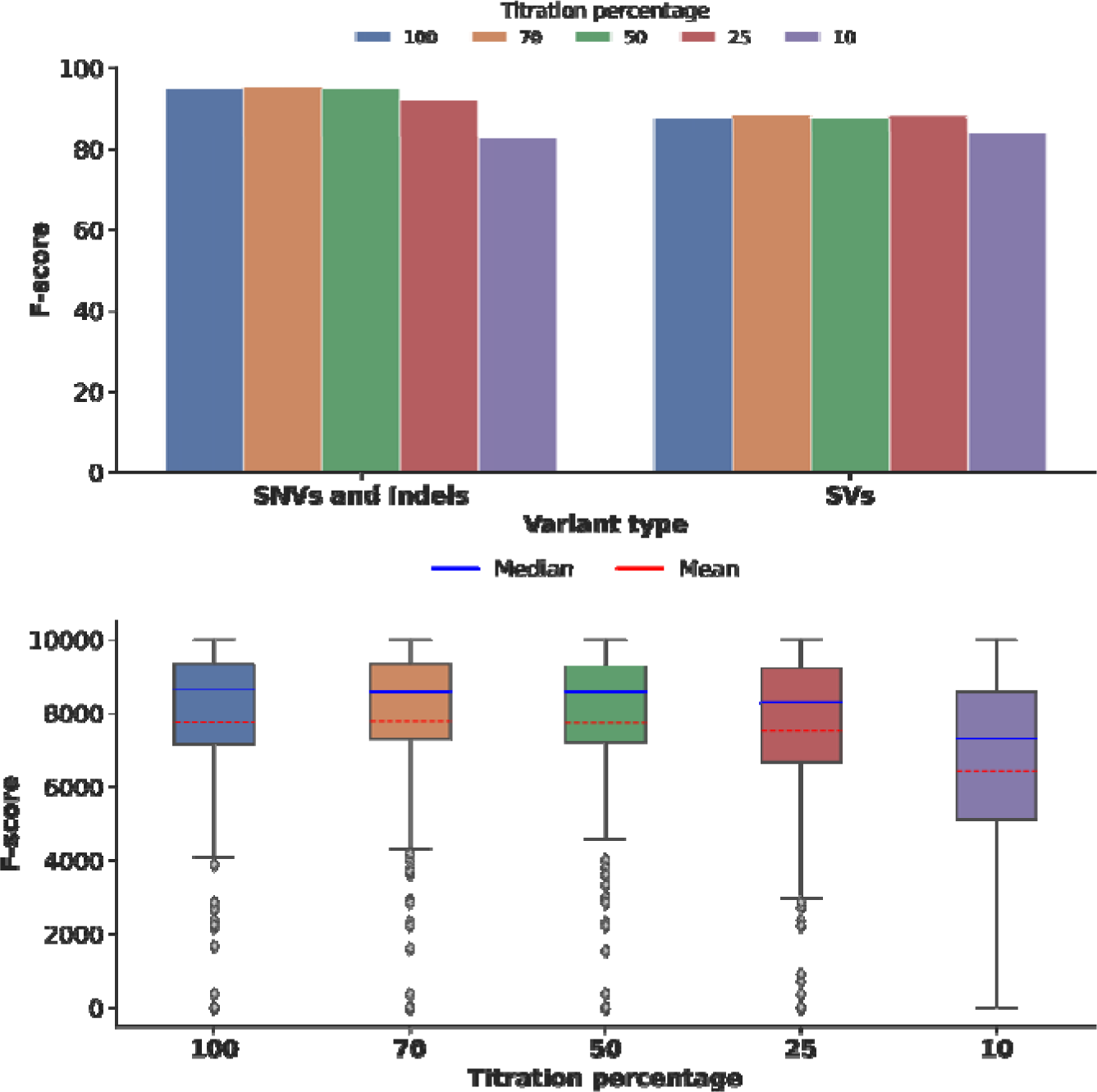
HG002 titration benchmark, sample HG002 coverage titration and its relation to the SNVs, Indels, and SVs accuracy. **a)** Compares SNV, indels, and SV F-scores across titration levels for sample HG002, with the variant type on the X-axis and the F-score percentage on the Y-axis. The titration values are color-coded: 100 in blue, 70 in orange, 50 in green, 25 in red, and 10 in purple. **b)** The boxplot illustrates the SNV F-score per gene across different titration levels. The X-axis represents the titration levels, while the Y-axis shows the F-score percentage. The solid blue line indicates the median, and the dashed red line represents the mean. This indicates that there is no performance difference while lowering the coverage and thus enables the increase of samples sequenced per run. Only at 10% of the original coverage (light green) we observe a performance difference.

### Systematic evaluation of TADGP across HG001 to HG007 cell lines

Building upon the performance of the TADGP panel with HG002 and its replicates, our next step was to explore how the panel and our analysis workflow would perform across 11 additional cell lines, which represent multiple human ethnic populations (HG001, HG003, HG004, HG005, HG006, HG007, HG1190, HG2723, NA12877, and NA12879). These samples include two trios (HG002-HG004 and HG005-HG007). An inherent challenge in this context is that genes may vary in the number of pseudogene copies, making the reference representation less accurate^50^. Thus, variant calling can become more challenging to represent genes correctly from one human population to the other. Furthermore, for many of these samples, there are no highly curated benchmarks available^39,51^.

We observed a high number of reads on target, as seen earlier for sample HG002 (**Supplementary Table S5**). The average per-gene coverage was 68.55x, with approximately 94.39% of bases at a depth greater than or equal to 8x (**Supplementary Figure 9**). There were, on average, 2.45 genes without coverage across the 11 samples (**Supplementary Figure 10** and **Table S5**). We benchmarked the panel’s performance against WGS data (Supplementary **Table S7**) as we did in the previous section. LR-WGS as well as SR-WGS exhibited different per-sample coverage, affecting the per-gene average coverage (**Supplementary Table S4**). The TADGP had better coverage compared to LR-WGS and SR-WGS, demonstrating the effectiveness of multiplexing samples on one SMRT cell.

We compared small variant calls between the TADGP, LR-WGS, and SR-WGS for samples HG001 to HG007 (excluding HG002). For this our analysis only includes 79.93% of these genes given limitations in GIAB variant benchmarks across these samples^52^, as illustrated in **Supplementary Figure 11**. The overall TADGP F-score for SNVs and indels ranged from 96.94% (HG007) to 98.41% (HG005) (**Supplementary Table S8)**.

Additionally, we analyzed the unique variants identified by each technology compared to the available benchmark set, as well as the variants that were shared across the different technologies. Across the six samples (HG001 to HG007), we observed that approximately 14,972.16 (66.35%) of the called variants are shared across all three approaches (TADGP, LR-WGS, and SR-WGS) and the benchmark set (**Supplementary Table S9**). Overall, there are on average 31 SNVs and indels per sample exclusively called by the panel and even missed by LR-WGS and SR-WGS. Furthermore, the panel reported fewer incorrect calls on average 548.66 per sample in contrast to e.g., Illumina reported 794.5 per sample. **Supplementary Figures 12-18** shows all the comparison details per sample for HG001-HG007. The Mendelian consistency for TADGP of small variants for both trios HG002 to HG004 and HG005 to HG007 was 94.94% and 96.19%, respectively (**Supplementary Table S11**). Likewise, the SV Mendelian consistency was high for HG002 (92.76%) and HG005 (94.68%). Furthermore, we conducted a phasing analysis across the 11 samples. Our results indicate that on average 95.94% of the genes were phased by TADGP with 23.43% of them within a single phase block (**Supplementary Table S10**). Overall, we observe that the TADGP performed well across the human population panel, discovering and phasing variants across medically relevant complex genes.

### Application of TADGP to unsolved rare disease cases

We used the TADGP on three trios, totaling nine samples, to investigate unsolved rare disease cases. These three families were chosen based on the suspicion of harboring causative variants within the TADGP. The samples encompass diverse ethnicities, including Asian and Hispanic, and present conditions including hypogammaglobulinemia, thrombocytopenia, lymphopenia, arthritis, and sclerosis (**Supplementary Table S2)**. These nine samples further underwent microarray and exon sequencing analysis without identification of the causative variant.

We run the samples in the TADGP to investigate whether it would be able to identify pathogenic variants in these families with unsolved rare diseases. Since DNA samples from those trios were extracted from blood and not from cell lines, we first assessed the coverage and read length distribution. We observed an average N50 read length of 6,189.83 bp with an 54.83% on-target rate, detailed information for each sample is provided in **Supplementary Table S4**. Additionally, we found that 96.40% and 92.71% of the bases exceed a coverage of 8x and 20x, respectively (**Supplementary Table S5)**. Our workflow identified on average 73,104.50 SNVs and 21,745.08 indels across the samples in addition to on average 1,413 SVs per sample (**Supplementary Table S12)**. The workflow was able to phase SNVs and SVs together (**Supplementary Table S10)**. Given these variant calls we measured the Mendelian consistency of the variants across the three families. We reported a high Mendelian consistency for SNVs and indels (96.36%) and SVs (94.36%) (**Supplementary Table S11**).

Next, we investigated potential *de novo* variants, which are often associated with sporadic inheritance disease patterns. For samples BH7648, BH9319, and BH9703, we observed 2, 1, and 3 *de novo* SNVs, respectively. We also explored SNVs that were homozygous in the proband but heterozygous in the parents, which could be deleterious (autosomal recessive inheritance: ARI)^53^. On average, we found that 3.29% of the SNV and indels to be ARI, see Supplementary **Table S11**. We annotated *de novo* and inherited homozygous SNVs and indels in the three families using Annovar^54^. Among *de novo* variants, we counted the number with Deleterious Annotation of Genetic Variants with Neural Networks (DANN) scores^55^, but none were predicted to be deleterious. Additionally, we identified 20 ARI (17 exonic, 2 intronic, and 1 in the UTR3) for sample BH7648 across 15 genes. Furthermore, for sample BH7648 we identified 13 exonic variants across 10 genes and for sample BH9703, 57 exonic and 7 intronic variants across 13 genes Supplementary **Table S11**. We suspected that specific genes might be associated with the phenotype of these progrants. A few of these genes are overlapping with the design of the TADGP, which include *IKZF1* (BH7648 and BH9703) and *CASP10* (BH9319) (**Supplementary Table S2**). In sample BH9703, we identified compound heterozygous exonic variants (**Figure 5a**), where a nonsynonymous paternal variant at position 50,368,335 A>G on chromosome seven is inherited in trans position with a maternal synonymous variant C>A at the same chromosome’s downstream position at 50,400,069. However, these two variants have allele frequencies of 0.7555 and 0.1633, respectively, in the gnomAD database. It might still be a trans effect as the frequency of both variants together is unclear. Further investigation will be needed to assess the potential functional impact of these variants.

**Figure 5.**
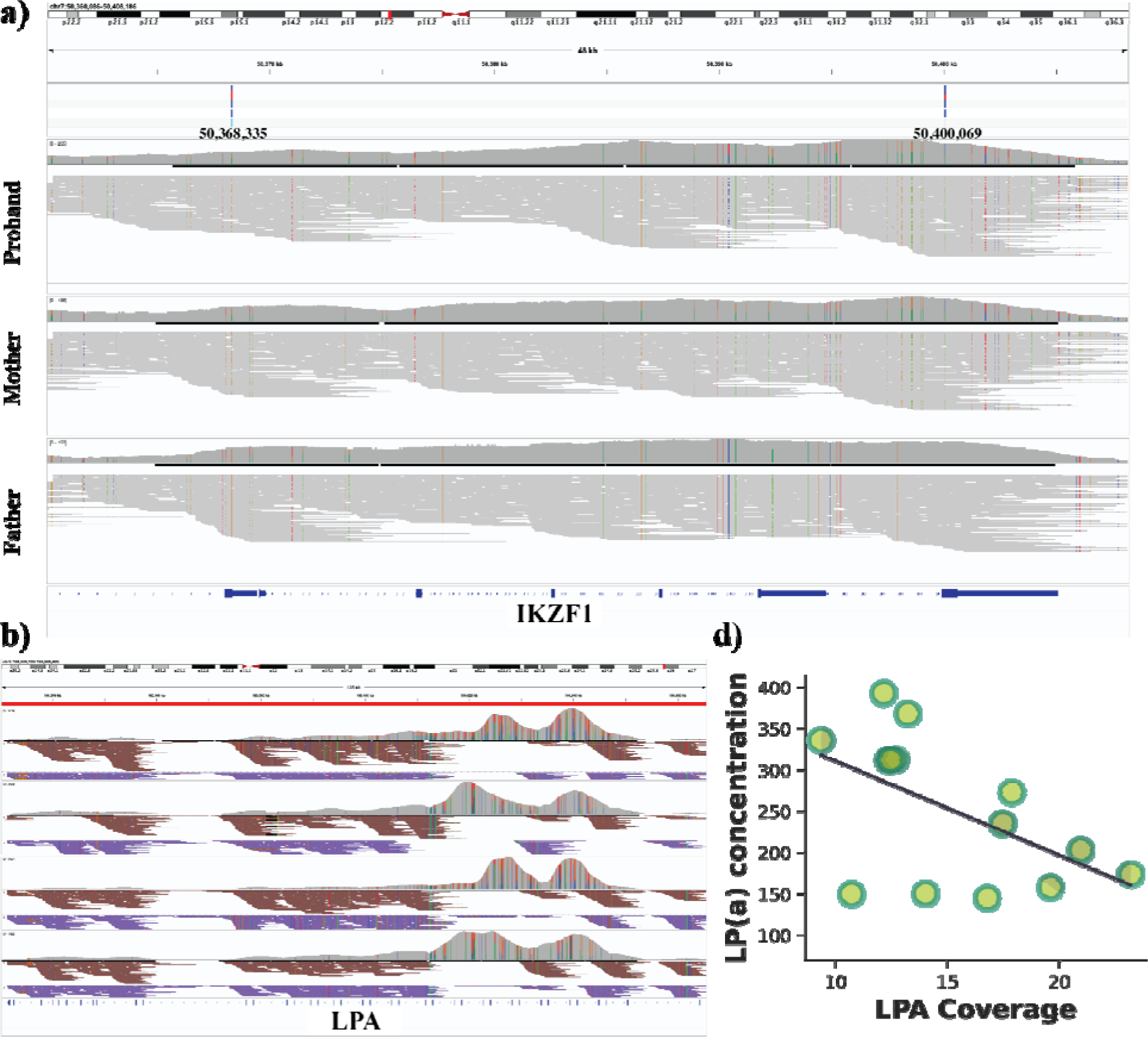
Application of TADGP on different rare and cardiovascular samples showing LPA copy number variant and its relation to lp(a) concentration. **a)** IGV screenshot displaying compound heterozygous variant in IKZF1 gene affecting the proband, which coils explain the condition, where a nonsynonymous paternal variant at position 50,368,335 A>G on chromosome seven is inherited in trans position with a maternal synonymous variant C>A at the same chromosome’s downstream position at 50,400,069. The three panels are proband, mother and father respectively. **b)** IGV screenshot displaying haplotype reads across four cardiovascular samples. Here the entire LPA gene was phased where reads are assigned to Hap1 (brown) and Hap2 (blue). The screenshot also highlights the KIV-2 copy number alteration with a distinctive double-humped pattern. **c)** A Scatter plot with a regression line showing the relation between LPA coverage on the X-axis and the concentration of lp(a) secretion on the Y-axis.

We did not identify any *de novo* SVs (details in **Supplementary Table S11**). Additionally, we analyzed SVs for homozygous SVs present in the proband but heterozygous in the parents (i.e., ARI). For samples BH7648, BH9319, and BH9703, we identified 42, 64, and 46 SVs, respectively and annotated the SVs using gnomAD^56^. We identified four ARI inherited SV in the intronic regions of BH7648 (*DMPK, DPP6, PADI4*, and *PDLIM3*), all categorized as benign or of unknown significance (**Supplementary Table S11**). Additionally, BH9319 harbored a benign intronic SV in *PDLIM3*, while two other SVs affecting *SEC63* and *PDSS1* introns were identified, one benign and one of unknown significance (**Supplementary Table S11**).

Thus, overall while the panel was not able to convincingly solve one of the three unsolved rare disease cases it highlighted multiple candidates that have not been observed before.

### Cardiovascular TADGP samples analysis

Given the success of TADGP to resolve complex genes and identify variants, we next applied it to a cohort of European and Hispanic descent investigating genomic based cardiovascular disease (CVD) risk. Here, we are especially interested in the complex gene *LPA* as it has a profound impact on Low-Density Lipoprotein (LDL) and thus CVD risk. *LPA* consists of multiple sections including KIV-2 that are highly polymorphic in the human population ranging from 5-50+ copies^24,57^. These copies include two exons and thus impact the overall length of mRNA and also expression of Lp(a) protein. Recent studies often rely on two SNVs (rs10455872 + rs3798220) to assess CVD risk as a proxy of the copy number variations^57^. Indeed, in Europeans these SNVs are in linkage with a high-risk copy number variant (CNV) of KIV-2. However, in other populations this association is often not the case^57,58^. We used TADGP across 14 samples of European (12 samples) and Hispanic (2 samples) populations where we also had LDL measurements.

All 14 samples showed consistent coverage with 90.82% of bases covered at minimum 20x (MQ20) (**Supplementary Table S4-5)**. On average, we identified 73,576.93 SNVs, 22,196.64 indels, and 1,392.79 SVs across these 14 patient samples (**Supplementary Table S12**). TADGP also phased on average 81.59% of the genes (**Supplementary Table S10).** We were interested in how well TADGP can be used to assess *LPA* specifically. We first manually inspected the *LPA* gene including the KIV-2 region (**Figure 5b**). Across these 4 randomly selected samples we can clearly see the duplication of the KIV-2 across the samples. Furthermore, we see that TADGP was able to phase reads in this region indicated by brown- and purple-colored reads (**Figure 5b**). The phasing of KIV-2 can be highly important as it might provide insights to different expression levels per haplotype. To quantify the ability of TADGP to recover the CNV state of KIV-2 we estimated the CNV number of this region by comparing it to a neighboring region within *LPA* (see methods for details). **Supplementary Table S2** provides the detailed CNV estimates. We correlated the CNV estimates with the obtained Lp(a) protein measurements. We observed as expected an inverse significant correlation (−0.53, p-value: 0.0493) of KIV-2 CNV and Lp(a) measurements (**Figure 5c**)^25^.

Besides *LPA* we investigated 70 other genes that are reported to impact CVD risk and are also captured by TADGP (**Supplementary Table S2**). We identified 2,293 (1.87%) of the variants across the exons of these genes. These exonic variants are observed in genes (*LPA*, *PDLIM3*, *CALR3*, *KCNE1*, *PDLIM3*, *CALR3)* linked to cardiac conditions based on ClinVar annotation. We further identified a systematic deletion of 82 bp in chromosome 22 (chr22:50,716,010) impacting *SHANK3* across 13/15 (86.66%) samples, including the control sample HG002. The SV genotypes across these samples were classified into 9 heterozygous variants (including HG002) and 4 homozygous variants. This specific deletion independent of the zygosity, is annotated to be related to Phelan-McDermid syndrome, which can cause cardiac problems. Interestingly, we identified this SV in only one individual within the gnomAD-SV database, suggesting its extreme rarity in the general population. However, it is unlikely that this deletion is indeed related to Phelan-McDermid syndrome as its prevalence is reported to be 2.5–10 per million births^59^. Thus, the lack of SV calling ability in these regions might lead to a misconception of the rarity of the particular deletion. Further analysis of the samples uncovered one sample exhibiting three deletions within intronic regions flanking exon 8 and 9 of *SHANK3*. Notably, previous studies in mice demonstrated that partial deletion of *SHANK3* exons 4-9 (*SHANK3* Δexon 4–9+/−) led to significant thickening of both the anterior and posterior walls of the left ventricle (LV), while complete deletion (*SHANK3* Δexon 4–9−/−) solely affected the anterior wall thickness compared to wild-type controls^60^. While additional functional validations are crucial to conclusively establish the impact of the identified variants, these findings show the potential of TADGP in capturing and analyzing notoriously challenging genes.

### Uniquely identified variants by the panel that evaded public db

Over this work, we demonstrated the reliability and utility of the TADGP in sequencing and analysis of 389 medically relevant yet challenging genes. To further investigate the significance of this approach, we examined how many of the variants discovered in this work are actually present in public databases. This could reflect the lack of knowledge about the diversity of these 389 genes and thus the importance of studying them to better understand what mutations might have an actual impact on these genes. To start this investigation, we started with HG002 to set the expectations. Here we actually first used the published SNVs and indels from GIAB directly and annotated them. First, to have a reference point, the genome wide SNVs and indels from GIAB overlapped to 99.16% with gnomAD directly. Next, for CMRG (273/389 genes) we saw that the percent of covered variants from gnomAD reduced to 97.66%.

Applying this analysis to our dataset of 34 samples (excluding the two HG002 replicates), we identified 155,653 variants (121,376 SNVs and 34,277 indels) present in at least four samples within the merged set. This threshold corresponds to an approximate minor allele frequency (MAF) of ≥12% across the entire dataset and thus clearly common variants. Notably, we were only able to annotate 80.59% of these common variants (89.22% of SNVs and 50.06% of indels) using the gnomAD database **Supplementary Table S14**. Interestingly, among the unannotated variants, exonic variants represented only 1.21% of SNVs and 0.19% of indels. For the 6,764 SNVs and indels that are shared across all samples, the number of variants that we can annotate increases to 82.82% (92.93% SNVs and 6.80% indels).

Next, we performed the same analysis for SV, which is harder to characterize^14,39^. Again, genome wide GIAB SV benchmark (v0.6) revealed only a 22.80% overlap with gnomAD-SV highlighting the larger challenge of capturing SV with short-read data in population databases. Extending this to the 217 GIAB CMRG SV benchmark, we did not find any SV annotated in gnomad-SV. For SV that are shared across a minimum of four samples in our collection (MAF

>12%) we only identified 7.99% of them in gnomAD-SV (**Supplementary Table S14)**. This highlights the need for improved annotation resources for SNV and SV calling across the 389 medically relevant genes. Even with the low number of samples we studied, here we could provide an annotation resource for future studies to provide limited but usable annotations in terms of population frequencies. We are thus releasing the alleles with population frequencies at: https://doi.org/10.5281/zenodo.10806570.

## Discussion

We introduced TADGP, a gene-targeted approach specifically designed for the cost-effective application of long-read sequencing in the investigation of 389 medically relevant genes known for their complexity. Throughout this study, we characterized the efficacy of TADGP across seven benchmark samples, including HG002 plus samples HG001 to HG007. In addition, we used the TADGP on a cohort of 23 patient samples, unveiling its efficacy in the identification and phasing of variants. Notably, this approach proved effective in the analysis of legacy samples (comprising 9 Mendelian cases) as well as blood samples (including 14 cardiovascular samples). TADGP consistently exhibited high coverage across all samples for the targeted genes and effectively captured other paralogous gene copies associated with the targeted genes. In conjunction with this targeted approach, we developed a new analytical workflow that integrates mapping, assembly, and targeted callers. This workflow aims to generate a harmonized and phased VCF file, simultaneously addressing both SNVs and SVs. We demonstrated a high accuracy and concordance across three replicates of HG002 GIAB samples. This accuracy was consistently observed across other samples and yielded new insights into the 389 medically relevant genes, even often outperforming whole genome sequencing approaches.

The design of TADGP is based on a previous study where we identified 386 challenging medically relevant genes in addition to 3 other genes (*GBA*, *CYP2D6*, and *SBDS*)^39^. This set of genes have been highlighted because of their repetitiveness and complexity of the regions they are located at. Moreover, reference errors in both GRCh38 and GRCh37 have been identified, posing obstacles to obtaining detailed characteristics for these genes^45^. The overall design across the autosomes includes 22.20 Mbp of sequence as we included the entire gene body along with the 3kb flanking region, which makes it possible to phase variants across the gene. To evaluate this, we used TADGP across 36 samples, with 12 samples per SMRT cell, resulting in high variant accuracy and phasing rates. To determine the cost benefit of TADGP, we conducted a subsampling experiment demonstrating the feasibility of using 48 samples per Revio SMRT cell, thus further reducing costs.

It should be noted that for a few genes the panel design did not work properly, whereas for four genes, the average sequence coverage was <1x in all our 36 samples. The overall assessment of coverage is occasionally complicated by the highly repetitive nature of several of these genes, including Variable Number Tandem Repeats (VNTRs).

Even with long reads, these challenging genes in the TADGP may not map well to the reference genome. To address this, we used a hybrid strategy to integrate mapping-based methods, targeted caller, and an assembly method to accurately identify variants. In certain cases, this hybrid strategy even outperforms whole genome sequencing.

TADGP can be used either independently or in conjunction with previously sequenced samples to enhance resolution across the 389 medically relevant genes. These genes were reported to exert a significant impact across diseases (**Figure 1a**).

However, only 25% of the genes have published ClinVar variants and thus show the need for more routine assessments of these medically important genes. This is also shown by the lack of gnomAD and gnomAD-SV annotations available for the variants we identified here. Only 80.59% of SNV and 7.99% of SV that are common across our samples (AF ≥ 12%) can be re-identified in gnomAD overall. It is surprising that numerous variants remain uncaptured by standard annotation databases such as gnomAD, underscoring the necessity for adopting the approach presented here to systematically evaluate variants within these complex medically relevant genes. To promote this, we provide the variants identified in this study as an annotation resource for further studies at https://doi.org/10.5281/zenodo.10806570.

TADGP establishes a cost-efficient and comprehensive approach for characterizing 389 medically relevant genes, thereby providing additional insights into multiple diseases and affected individuals on a large-scale.

## Methods

For the rare disease cases, all individuals provided consent under the BH-CMG (GREGoR) IRB protocol H-29697. Regarding the cardiovascular samples, the study was approved by the Baylor College of Medicine (BCM) Institutional Review Boards (protocol number: H-43884). Written informed consent was obtained from all study participants.

### Capture panel description

In brief, double-stranded DNA probes were designed to GRCh38 regions using a sparse tiling approach which is supplemented by a repeat screen and GC content optimization. The panel was further optimized by shifting and adding probes to poor-performing regions based on the first round of experiments. The total target size covered is 22.20 Mbp with 24,363 probes. A total of 389 regions were targeted, except *LCE3B, SNORD64, DUX4L1, and TRBV9*, which were not covered due to high off-target rate. We used a probe length of 120 bp.

### Pipeline description

We built our analysis workflow using snakemake v7.22.0^61^ running on a python v3.9 interpreter. Multiplexed samples were first demultiplexed and adapters were trimmed using lima (https://github.com/PacificBiosciences/barcoding) v2.5.0.

For each sample, HiFi reads were marked for PCR duplicates with pbmarkdup v1.0.2 (https://github.com/PacificBiosciences/pbmarkdup) and aligned with pbmm2 v1.7 (https://github.com/PacificBiosciences/pbmm2) to the reference using GRCh38 fixed reference. Aligned reads were further processed through DeepVariant v1.5.0^46^ and Whatshap^62^ v1.1 for SNV calling and phasing of reads and variants, respectively. Structural variants were called from haplotagged reads by sniffles^63^ v2.0.5.

In a separate pathway, HiFi reads were assembled using hifiasm^64^ v0.15, followed by alignment to the reference using minimap2^65^ v2.17. Variants of all sizes were called from the aligned assemblies using htsbox (https://github.com/lh3/htsbox) build r346.

After all variants were called from aligned reads and assemblies, we used bcftools^66^ v1.13 to annotate each intermediate VCF file according to calling source by including the INFO/V field as indicated by the appended header lines:

##INFO=<ID=GENE,Number=1,Type=String,Description=“Gene Name”>

##INFO=<ID=V,Number=1,Type=Integer,Description=“V=1 Assembly V=2 HiFi_Reads”>

Labeled variants were filtered according to the user input defined in the target BED (see below) and combined to form the final output VCF files.

Input to the workflow includes a target BED file with 6 columns: CHROM, FROM, TO, GENE, SNV, SV. The “gene” column is a unique target name, typically the gene, while the last two columns define which variant-calling pathway to take for SNV and SV, respectively. A value of 1 indicates variants should be sourced from the aligned assembly, and a value of 2 indicates variants sourced from aligned HiFi reads (see also INFO fields above). The workflow generates calls for both variant classes for all targets using both mapped HiFi reads and assemblies, and then filters and combines variant calls based on the BED. Output SNV VCF files contain all variants for which the INFO/GENE and INFO/V values correspond to the GENE, SNV columns of the BED, while the output SV VCF contains variants with values according to columns GENE, SV as described above.

### Sample DNA extraction and sequencing

Genomic DNA (gDNA) samples were obtained from the Coriell Institute. Around 500-1000 ng of gDNA were sheared using Covaris g-TUBE. Fragmented gDNA were subjected to the <u>Twist</u> <u>Long-Read Capture Protocol</u>. After end-repair and a-tailing, truncated Y-shaped adapters were ligated onto the adapted gDNA. A pair of 10-bp unique dual indices (UDIs) for sample barcoding were added during PCR. 4-8 samples were pooled in a single tube for overnight hybridization. A custom panel (now manufactured as <u>Twist Alliance Dark Genes Panel</u>) from Twist Bioscience was used to perform targeted capture of the regions of interest. The post-capture libraries then underwent SMRTbell library preparation using SMRTbell® prep kit 3.0 and sequencing on a PacBio Revio instrument (150pM, 24-hour movie) according to the manufacturer’s protocol using the High Fidelity (HiFi) sequencing protocol^67^. Up to 4 samples were multiplexed and sequenced in one Sequel SMRT Cell or 12 samples in one Revio SMRT Cell with HiFi read length of 5-10 kb.

The data generated underwent initial processing onboard the Sequel IIe/Revio instrument, using the PacBio SMRT Link software. Subsequently, the onboard analysis included base calling, HiFi read generation Following this stage, the files are transmitted to the HGSC compute cluster for further post-processing.

### Variant calling for PacBio WGS

For SNVs, indels, and SVs calling and phasing we used PRINCESS^68^ version 2 with the default parameters and subcommand all, with the ′--ReadTypè set to ′ccs′.

### SNVs and SVs calling for Illumina

For Illumina whole genome sequencing, we aligned the reads using BWA-MEM^69^ version 0.7.17-r1188 with default parameters. Subsequently, SNVs and indels were called using the GATK HaplotypeCaller^70^ version 3.6-0-g89b7209 workflow, and for structural variants, we used Manta version 1.6.0 with default parameters.

### Benchmarking SNVs and SVs

We conducted SNV benchmarking separately by using the gene coordinates as input for RTG vcfeval version 3.12.1 (https://github.com/RealTimeGenomics/rtg-tools) Supplementary section “QC Pipeline’’. For benchmarking the entire gene set, we followed the same process. The benchmarking process was carried out using the default parameters of RTG vcfeval. The input BED regions and SNPs were sourced from GIAB CMRG version 1.0^52^ (https://ftp-trace.ncbi.nlm.nih.gov/giab/ftp/release/AshkenazimTrio/HG002_NA24385_son/CMRG_v1.00/GRCh38/SmallVariant/). For samples HG001, HG003, HG004, HG005, HG006, and HG007, we used GIAB data available at https://ftp-trace.ncbi.nlm.nih.gov/giab/ftp/release/. To ensure accuracy, we filtered both the BED regions and SNV regions using ′bedtools intersect′^71^, with the input being the BED regions from CMRG. To benchmark SVs, we used ′truvari bench′ version 3.2.0^72^ with the default parameters and used the following flags: ′--passonly′, ′--pctsim=0′, and ′--multimatch′. We used a BED input file input and an SVs VCF file sourced from CMRG consortia version 1.0.

To benchmark genes that were not available from GIAB, we first selected them using the ′bcftools intersect′ tool with the ′-v′ option. The ′-à argument was set to the 389 genes, and ′-b′ was set to the 273 genes available from the GIAB consortium. Next, we used the variants called by HTSbox from the HG002 assembly (accession ID PRJNA727430) and intersected them with the BED file for the 116 genes. We filtered out multiallelic variants using the ′bcftools view -M2′ option and indexed the variants with ′tabix′ version 1.12^73^.

To validate variant calls made by Paraphase in S*MN1/SMN2, PMS2/PMS2CL, NCF1/NCF1B/NCF1C* and *CFC1/CFC1B*, we compared per haplotype VCF files against the HG002 T2T assembly v1.0 (https://github.com/marbl/HG002) (variants called by HTSbox).

### QC of the workflow results

We used nanoplot^74^ version 1.40.0 with default parameters and included the flags ′--N50 -- alength --no_supplementary′ to calculate whole genome coverage. For normalized coverage, we divided the average gene coverage by the maximum average gene coverage, thereby scaling all coverages to a maximum of one. To identify on-target coverage, we used samtools^75^ (version 1.12). Initially, we determined the total reads in the aligned BAM file using the command ′samtools view -c -F 3076′.

Subsequently, we ascertained the number of reads on target using the same command but after extracting the on-target reads with a buffer of 10,000 bp. To achieve this buffer, we used bedtools (version 2.30.0) with the command ′bedtools slop -b 10000′. The intersections were computed using ′bedtools intersect′.

To determine the base pairs on target, we used samtools and AWK. This involved extracting reads with the flag ′3076′ and then summing the lengths of the 10th field. The same procedure was applied to the on-target region. Average gene coverage was calculated using mosdepth^76^ (version 0.3.2) with the ′--by′ option to support gene coordinates and a ′--mapq 20′ threshold.

To calculate the bases with 8x, 10x, and 20x coverage, we used the per-base coverage output from mosdepth along with bedtools and AWK Supplementary section “Gene coverage”. Initially, we intersected the per-base coverage with gene coordinates using ′bedtools intersect′. Subsequently, we used AWK to sum the values that exceeded 8x, 10x, and 20x coverage.

For the whole genome sequencing (WGS) data from both Illumina and HiFi, we determined genome coverage using nanoplot^77^. This involved dividing the total bases aligned by ′3,088,286,401′, which is the estimated count of the human genome bases. The method for calculating average gene coverage was identical to that used for the panel, as explained earlier.

### Pseudogene identification

We expanded the gene regions by one kilobase using ′bcftools slopè, followed by selecting the sequence for each gene using ′bedtools getfastà. Subsequently, we masked the genome using ′bedtools maskfastà and aligned the gene fasta file to the masked genome using ′minimap2′ with the ′-c′ flag to incorporate the ′CIGAR′ number. From the resulting PAF file, we extracted aligned genes with a minimum alignment block length of 2 kilobases and calculated the identity by dividing the number of residue matches by the query sequence length. We retained only those genes with an identity of at least 0.90. Finally, we identified the genomic regions to which these genes align and utilized them to compute coverage during the alignment of the total reads, both with and without the fixed GRCh38 reference, using ′mosdepth′ with ′by′ set to the genomic region the genes aligned to.

### Sample titration

To generate titrations representing 70%, 50%, 25%, and 10% of the total run from the multiplexed BAM file, we used ′samtools view′. The ′-s′ parameter was assigned the values of ′11.7′, ′11.5′, ′11.25′, and ′11.10′ to perform this titration process, where the see set to ′11′.

### Merging and identifying unique variants

We merged the SNVs and indels using ′bcftools merge -Oz -ò, combining the data from the 34 samples obtained in the three SMRT cell runs. We selected variant that are shared with at lear 4 samples by using ′bcftools view -i ‘count(GT!=“mis”)>3’′.

Regarding SVs, we used SURVIVOR merge with the following parameters: SVs_merge.txt 1000 0 1 0 0 50.

- SVs_merge.txt denotes the file containing paths to the input SVs files
- 1000 bp represents the maximum distance between two SVs
- The maximum number of supporting callers is set to 0
- SV type is considered during merging
- Strand information is disregarded
- And distance estimation based on SV length is not conducted, and the minimum SV size is set to 50.

To select SVs that are shared within at least four samples we used ′bcftools view -i ‘SUPP!=“1” && SUPP!=“2” && SUPP!=“3” ′.

### Number of genes that intersects with variants from ClinVar

We used the ClinVar VCF file available at https://ftp.ncbi.nlm.nih.gov/pub/clinvar/vcf_GRCh38/archive_2.0/2023/clinvar_20230107.vcf.gz. Subsequently, we filtered variants using bcftools with the following criteria: ′bcftools view -i ‘CLNSIG=“Pathogenic” & CLNREVSTAT=“criteria_provided” & CLNREVSTAT=“_multiple_submitters” & CLNREVSTAT=“_no_conflicts” & CHROM!=“X” & CHROM !=“MT” & CHROM!=“Y”’ clinvar_20230107.vcf.gz′. This filtering retained only pathogenic variants where the criteria were provided, supported by multiple submitters, and had no conflicts. Additionally, we excluded mitochondrial variants as well as those located on chromosome X and Y.

### Variants annotation

We annotated SNVs and indels using Annovar^54^ latest version table_annovar.pl with reference build hg38 and the following protocols: gnomad312_genome (20221228), dbnsfp30a, clinvar_20221231. Additionally, we applied filters f, f, f for these annotations. For SVs, we used SVAFotate version 0.0.1 with gnomAD database annotation from the SVAFotate GitHub repository SVAFotate_core_SV_popAFs.GRCh38.bed.gz, which used gnomAD-SV_v2.1 and AnnotSV^78^ version 3.3.6.

### CNV detection in KIV exon II

To compute the ratio between KIV-2 coverage and other regions within the *LPA* gene, we designated the region chr6:160,638,526-160,639,526 (GRCh38) as the KIV-2 region and the region chr6:160,531,482-160,532,482 as the control region. using samtools view, we tallied the number of reads in each region. Subsequently, we calculated the ratio by dividing the read counts from the KIV-2 region by those from the control *LPA* region.

### Identifying variants intersecting with suspected genes for rare sample cases

We intersected a list of suggested genes, which could potentially explain some of the diseases in the Mendelian rare cases. The gene coordinates were obtained from the Biomart website for human genome GRCh38, and we carried out this operation using bedtools version 2.30.0 with the default ′intersect′ options.

### Mendelian consistency for SNVs and SVs

For Mendelian consistency assessment of both SNVs and SVs, we used the bcftools plugin +mendelian with default parameters, and specified the relationship using the -t flag.

## Data availability

Supplementary Table S7 lists the raw reads were downloaded from SRA The GIAB benchmark is downloadable here:

HG001:

SNVs and Indels: https://ftp-trace.ncbi.nlm.nih.gov/ReferenceSamples/giab/release/NA12878_HG001/latest/GRCh38/

HG002:

SNVs and Indels: https://ftp-trace.ncbi.nlm.nih.gov/ReferenceSamples/giab/release/AshkenazimTrio/HG002_NA24385_son/CMRG_v1.00/GRCh38/SmallVariant/

SVs: https://ftp-trace.ncbi.nlm.nih.gov/ReferenceSamples/giab/release/AshkenazimTrio/HG002_NA24385_son/CMRG_v1.00/GRCh38/StructuralVariant/

HG003:

SNVs: https://ftp-trace.ncbi.nlm.nih.gov/ReferenceSamples/giab/release/AshkenazimTrio/HG003_NA24149_father/latest/GRCh38/

HG004:

SNVs: https://ftp-trace.ncbi.nlm.nih.gov/ReferenceSamples/giab/release/AshkenazimTrio/HG004_NA24143_mother/latest/GRCh38/

HG005:

SNVs: https://ftp-trace.ncbi.nlm.nih.gov/ReferenceSamples/giab/release/ChineseTrio/HG005_NA24631_son/latest/GRCh38/

HG006:

SNVs: https://ftp-trace.ncbi.nlm.nih.gov/ReferenceSamples/giab/release/ChineseTrio/HG006_NA24694_father/latest/GRCh38/

HG007:

SNVs: https://ftp-trace.ncbi.nlm.nih.gov/ReferenceSamples/giab/release/ChineseTrio/HG007_NA24695_mother/latest/GRCh38/

ClinVar VCF file is available here: https://ftp.ncbi.nlm.nih.gov/pub/clinvar/vcf_GRCh38/archive_2.0/2023/clinvar_20230107.vcf.gz

Variant calling VCF files are available at: https://doi.org/10.5281/zenodo.10806570

## Code availability

The pipeline is available at https://github.com/PacificBiosciences/HiFiTargetEnrichment

The QC pipeline for the panel is available at https://github.com/MeHelmy/HiFiTargetEnrichmentQC

## Acknowledgment

FJS and MM are supported by NIH grants (1U01HG011758-01).

SIH S10 grant 1S10OD028587.

We would like to thank Luis F. Paulin for his fruitful discussion on the *de novo* SVs calling. We would like to thank Barbara Bock for her help on gene annotations.

## Authors contribution

FS designed the study. MM and JH analyzed the data. FS, ZK, and ME provided critical feedback and oversaw the project. HC, TH, CR, TF designed the probes for the panel. TH, HC, SJ, HD, QM, CL, SZ, PB, TH, YH, GH, SK, and DM optimizing the sequencing of the panel. MM, JH, XC, ZK, FS, and ME leading the development of the analysis software and the analysis of the data. All authors contributed to writing the manuscript.

